# Likelihood of blood culture positivity using SeptiCyte RAPID

**DOI:** 10.1101/2025.05.09.25327025

**Authors:** Krupa Navalkar, Alyse Wheelock, Melissa Gregory, Danielle Clark, Hannah Kibuuka, Stephen Okello, Sharon Atukunda, Abdullah Wailagala, Peter Waitt, Francis Kakooza, George Oduro, Nehkonti Adams, Maximilian Dietrich, Maik von der Forst, Marcus J. Schultz, Neil R. Aggarwal, Jared A. Greenberg, Silvia Cermelli, Thomas D. Yager, Richard B. Brandon

## Abstract

Early diagnosis and identification of causative pathogens using blood culture in patients suspected of Blood Stream Infection (BSI) and sepsis are critical for improving patient outcomes through early and more targeted treatment. There is a need for tools that can guide the use of microbiologic diagnostics, especially where resources are limited, such as in lower and middle income countries (LMICs), pandemic and mass-casualty scenarios, and prolonged field care settings during military operations.

**Methods:** Post-hoc retrospective analysis of individual patient data from three prospective clinical studies, conducted in North America, Europe and Africa, to investigate the association between SeptiCyte RAPID test results (SeptiScores) and blood culture (BC) results.

**Hypothesis:** that a significant correlation exists between elevated SeptiScores and positive blood culture results, and between low SeptiScores and negative blood culture results.

**Results:** The area under the receiver operating characteristic curve (ROC AUC) was 0.91 for 85 BC(+) versus 257 SIRS, and was 0.80 for 164 BC(-) versus 257 SIRS. As the SeptiScore increases, the relative probability of a septic patient being BC(+) as opposed to BC(-) also increases. A non-linear positive correlation is observed. Below a crossover point at SeptiScore 10, the ratio of probabilities of BC(+) sepsis / BC(-) sepsis is <1 while above the crossover point this ratio is >1. Thus, septic patients with SeptiScores >10 have a higher probability of being BC(+) compared to BC(-).

**Conclusions:** Elevated SeptiScores, obtained before blood culture results, are indicative of increased blood culture positivity. This may have clinical utility, particularly in resource limited settings, as an aid for improving the efficiency of blood culture practice, for instance by informing patient selection and interpretation of blood culture results.

## 1. Introduction

Bloodstream infections (BSI) are a major cause of morbidity and mortality in both hospital and community settings worldwide [1–3]. Early diagnosis and identification of causative pathogens in patients suspected of BSI and sepsis are critical for improving patient outcomes through early and more targeted treatment. This unmet need is most acute in poorly resourced environments, which include not just healthcare settings throughout the lower and middle income countries (LMICs) [4, 5], but also pandemic [6, 7] and mass-casualty scenarios [8–10], and prolonged field care settings during military operations [11, 12].

A lack of early BSI diagnosis has led to more patients receiving inappropriate empirical therapy and to an increase in overall mortality and antimicrobial resistance [13]. Diagnosis of BSI largely relies on culture-based methods that have evolved little in decades [14]. Some challenges associated with traditional culture-based methods for diagnosing BSI include lack of timeliness [15], reduced sensitivity associated with prior antibiotic use (false negatives), and contamination (false positives).

Empirical data indicates that >90% of all blood cultures (BC) taken are negative [16–18], and only ∼50% of patients retrospectively diagnosed with sepsis have positive blood culture results [19]. Of those blood cultures that are positive, a contamination frequency as high as 16-23% has been reported [18]. Blood culture contamination is known to lead to increased pharmacy and laboratory costs, increased length of hospital stays, and unnecessary antibiotic use [18, 20]. In addition, the interpretation of blood culture results (pathogen or contaminant) is not always straightforward, and may depend on the background and experience of the clinician [21–25]. These clinical conundrums are often even more complex in the context of war-related polytrauma and burn injuries [10].

Thus, shortcomings in clinical practice related to testing for BSI are evident and have been noted previously [25]. Various recommendations have been proposed aimed at decreasing the frequency of unnecessary blood cultures, for taking blood cultures of optimum volumes at optimum times, and for optimized phlebotomy techniques to decrease the frequency of contaminants [26]. The suggestion has also been made to take blood cultures only when certain leading indicators are positive [Fabre et al. 2020] [27]. On this point, temperature spikes alone appear to be insufficient as a predictor of when blood cultures should be taken [Riedel 2008 and references therein] [28 and references therein].

SeptiCyte RAPID is an FDA-cleared, host-immune-response test with a one-hour turnaround time that provides a likelihood of sepsis on a scale of 0 - 15 (SeptiScore) (Balk et al., 2024a, 2024b). [29, 30] The test is based on quantitative analysis of the ratio of mRNA expression levels of two single-copy genes, PLAC8 and PLA2G7. These genes were first identified as informative for sepsis diagnosis through an unbiased bioinformatic screen of microarray data that did not incorporate any assumptions about their biological function [McHugh et al., 2015] [31] PLAC8 (also known as Onzin) is a small, highly conserved protein involved in regulating cell differentiation, innate immune responses, and autophagy. It has been linked to functions in placental development as well as roles in cancer cell proliferation and metabolic stress responses [32]. PLA2G7 encodes lipoprotein-associated phospholipase A2 (Lp-PLA2), an enzyme that hydrolyzes oxidized phospholipids in LDL particles to produce pro-inflammatory mediators. It plays key roles in vascular inflammation and has been widely studied as a biomarker and potential therapeutic target in cardiovascular disease [33]. The specific roles of these two genes in biological processes directly linked to sepsis is still under investigation in multiple laboratories (e.g. Zhang et al., 2024; Jin et al. 2024). [34, 35].

We hypothesized that a significant correlation exists between elevated SeptiScores and positive blood culture results, and between low SeptiScores and negative blood culture results. To evaluate this hypothesis, we conducted a retrospective statistical analysis of data from three independent observational clinical studies. While recognizing that such correlations may be imperfect—and that low SeptiScores should not provide unwarranted reassurance in isolation—data supporting the hypothesis would nonetheless suggest that SeptiScores could help improve the efficiency of blood culture practice, for example by informing patient selection for initial or repeat blood culturing.

## 2. Materials and Methods

### 2.1. Study Cohorts

Patients were derived from three independent obervational clinical studies, herein called “510k”, “SeptAsTERS” and “UGANDA”. The design of these clinical studies has been described in detail elsewhere (respectively Miller et al., 2018; von der Forst et al., 2024; Blair et al., 2023) [36–38]. **Figure 1** shows a flow diagram that describes the selection of patients included in this study.

**Figure 1.**
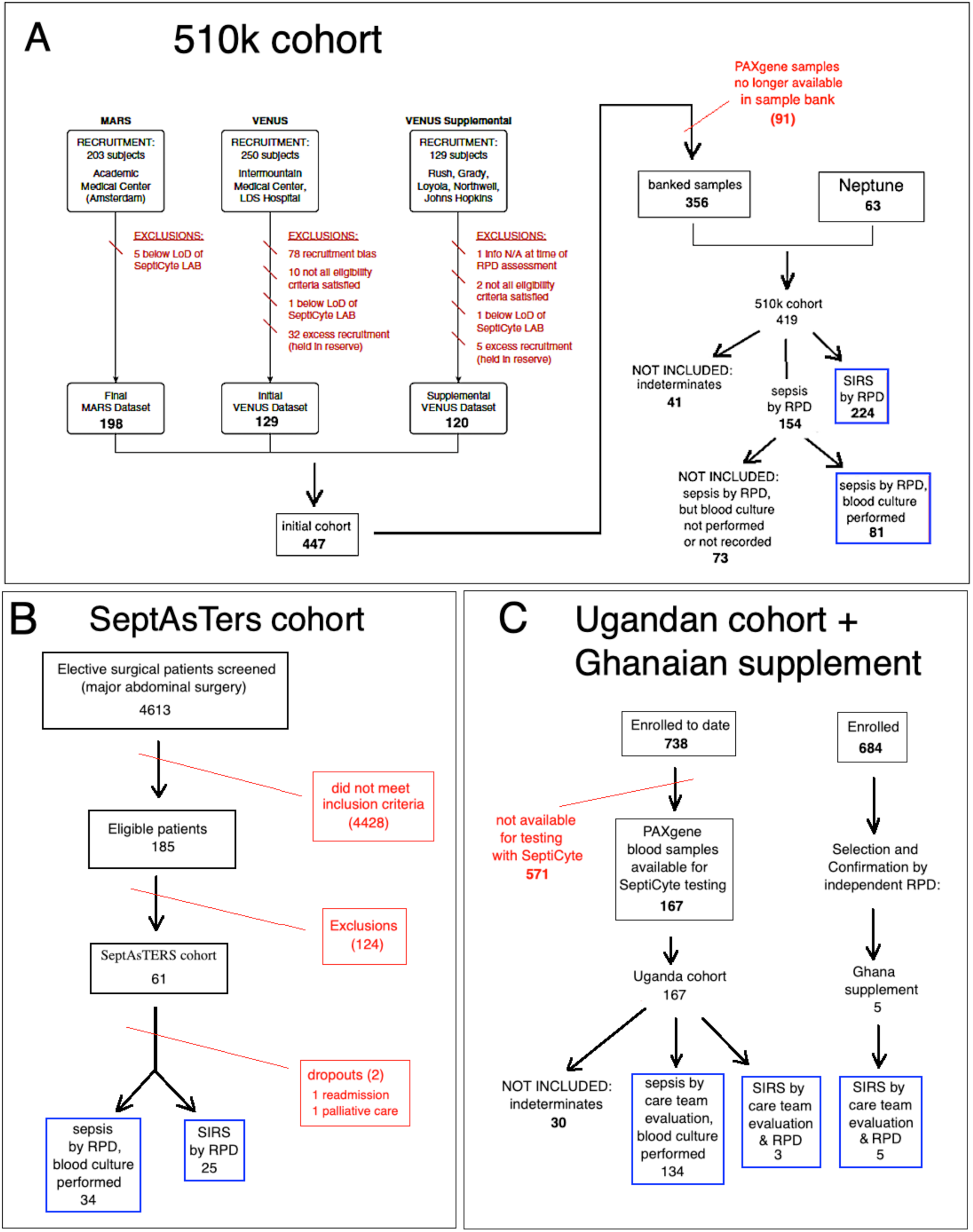
Flow Diagram of Patients Included in This Study. The initial pool from which the final patient selection was made consisted of 652 patients from three prior studies. The final pool consisted of subjects from (A) the 510k cohort, (B) the SeptAsTERS cohort, and (C) the Ugandan cohort + Ghanaian supplement. Patients were excluded from consideration for either of three reasons: 1) if they fell under protocol exclusion criteria; 2) were deemed “indeterminate” (i.e. could not be confidently diagnosed as either SIRS or sepsis; or 3) if they were diagnosed as sepsis but blood cultures were either not performed or not recorded. Details are indicated in the figure.

Patient classification as “sepsis” versus “SIRS” was determined using one of two approaches, depending on study site and available resources. **Method 1:** In the 510k and SeptAsTERS studies, sepsis and non-infectious systemic inflammatory response syndrome (SIRS) were adjudicated via Retrospective Physician Diagnosis (RPD) [36], in which an external panel of physicians, independent of patient care, reviewed clinical data. This method aimed to minimize inter-site bias and to standardize diagnostic criteria, but may be constrained by incomplete documentation, absence of bedside context, and variability among panelists. **Method 2:** In the Ugandan study, diagnoses were based on attending physicians’ assessments at multiple time points. This approach leveraged direct clinical knowledge of patients which enabled detection of subtle features, although it carried a greater risk of site-specific bias.

#### 2.1.1 The “510k” study

The “510k” study, conducted at multiple sites in the USA and Europe, enrolled a total of 419 patients from the intensive care unit (ICU) who displayed SIRS and were suspected of sepsis. By consensus RPD, 154 (36.8%) of these patients were determined to have sepsis, and 224 (53.5%) were determined to have SIRS. There were 41 patients (9.8% of total) who could not be classified unambiguously as sepsis or SIRS. These were considered “indeterminates” and are not included in the present analysis. Blood cultures were ordered on the basis of clinical judgement and accordingly were only taken if deemed necessary by the care team. Of the 154 patients classified as septic, 81 had blood cultures taken, and of these 48 (59.3%) were blood culture positive (see Figure 1). The comparator for classification as SIRS or sepsis in this study was consensus clinical adjudication by an external 3-member panel of expert clinicians. The adjudication process has been described in detail in the online data supplement Part 3, in Miller et al., 2018 [36]. The RPD process generally conformed to the conceptual framework provided by the Sepsis-2 definition (Levy et al., 2003) [39]. The adjudicators did not have access to the SeptiCyte RAPID test results.

#### 2.1.2 The “SeptAsTERS” study

The “SeptAsTERS” study, conducted at a single hospital in Germany, enrolled 57 postsurgical ICU patients with SIRS and showing signs of clinical deterioration. A total of 32 (56.1%) were retrospectively determined to have sepsis, and 25 (43.9%) were determined to have SIRS (see Table 1). Blood cultures were ordered on the basis of clinical judgement and accordingly were only taken if deemed necessary by the care team. Of the 32 septic patients, 15 (46.8%) were blood culture positive. The comparator for classification as SIRS or sepsis in this study was a post-hoc assessment of all patients by three independent intensive care professionals, who were not involved in the study. The process followed a model similar to the one described in the online data supplement Part 3 by Miller et al. [2018] [36] based on FDA Guidance and on publications by Klein Klouwenberg et al. [40,41]. Details of the post-hoc assessment process are provided in von der Forst et al. (2024) [32]. The RPD process adhered largely to the conceptual framework of the Sepsis-2 definition (Levy et al., 2003) [39]. Nevertheless, the incorporation of organ-dysfunction criteria means that the resulting assessments also shared features with the Sepsis-3 framework [42]. The adjudicators did not have access to the SeptiCyte RAPID test results.

**Table 1.** Patient Demographics and Clinical Parameters. Abbreviations: SOFA, sequential organ failure assessment (score); M, male, F, female; ND, not determined. A two-sample proportions test was used to estimate p-values for the sex ratio in each study. Pearson’s chi-squared test was used to estimate the p-value for comparing SOFA score distributions between the three studies.

| Patient characteristic | Clinical Study |  |  | p-value |
| --- | --- | --- | --- | --- |
|  | 510k | UGANDA | SeptAsTERS |  |
| Total number | 305 | 137 * | 59 | ND |
| Age, median (range) | 58 (18-90) | 46 (18-86)<br>[missing: 12] | 66 (30-84) | 0.072 |
| Sex (M/F) | 172 (56%) M<br>133 (44%) F<br>(p = 0.037) | 65 (47%) M<br>72 (53%) F | 38 (64%) M<br>21 (36%) F<br>(p = 0.038) | 0.064 |
| Race / Ethnicity |  |  |  | p < 0.00001 |
| White | 189 (61.9%) | 0 | 59 (100%) |  |
| African or African heritage | 82 (27%) | 137 (100%) | 0 |  |
| Asian | 14 (4.6%) | 0 | 0 |  |
| Hispanic | 16 (5.2%) | 0 | 0 |  |
| Other/Unknown | 4 (1.3%) | 0 | 0 |  |
| SOFA, median (observed range)<br>[number and % missing values] | 5 (0-20)<br>[missing values: 34/305 = 11.1%] | 2 (1-10)<br>[no missing values] | 5 (1-18)<br>[no missing values] | <0.001 |
| Sepsis | 81 <sup>1</sup> | 134 <sup>2</sup> | 34 <sup>1</sup> | ND |
| SIRS | 224 | 3 * | 25 | ND |
| <sup>1</sup> Patients classified as septic by consensus RPD, for whom blood culture results were available<br><sup>2</sup> Patients classified as sepsis / severe sepsis / septic shock by the on-site care team at multiple timepoints during the patient's hospital stay (screening, 0h, 6h, 24h and in some cases 72 h). Data included from 0h timepoint only.<br>* 5 SIRS patients from the Ghanaian cohort are not included in this table, because only aggregate clinical data was available. See Supplement S1. |  |  |  |  |

#### 2.1.3 The “UGANDA” study

The “UGANDA” study, performed at Fort Portal Regional Referral Hospital, Kabarole District, Western Uganda and ACESO Uganda (Makerere University Walter Reed Program and Infectious Disease Institute), Kampala, Uganda, comprises a subset of a larger study which is described in Blair et al. (2023) [38]. The 167 adult patients considered here presented initially to hospital with clinical signs of SIRS and a suspicion of sepsis, and ultimately were recruited from the emergency department (ED) and from medical wards (as opposed to the ICU as was the case for patients in the 510k and SeptAsTERS studies). Patients were excluded if aged <18, weighed less than 40kg, or were on chemotherapy, asplenic, immunosuppressed or pregnant. As dictated by protocol, blood cultures were taken for every patient. The comparator for classification as SIRS or sepsis in this study was based on the sepsis-2 criteria (Levy et al., 2003) [39]. Patient evaluation was conducted by the on-site care team at multiple timepoints (screening, 0h, 6h, 24h, and in some cases 72h). Of the 167 patients enrolled in this study, 134 (80.2%) were evaluated as sepsis / severe sepsis at presentation, and 3 (1.8%), although evaluated as sepsis/severe sepsis at presentation, were later retrospectively adjudicated as SIRS by an independent clinician. A total of 30 patients (18.0%) could not be classified as SIRS or sepsis, and therefore were designated “indeterminate” and not included in subsequent analyses. The adjudicators did not have access to the SeptiCyte RAPID test results.

#### 2.1.4 The Ghanian supplement

To supplement the number of SIRS patients from Uganda and to include SIRS cases from a distinct African site, five additional SIRS patients from Ghana were added to the study.

These patients were selected from “discovery cohort” participants in “An Observational Study of Sepsis in Kumasi, Ghana”, protocol number NMRC.2016.0004-GHA [43,44]. These patients had previously been adjudicated under the Sepsis-2 framework by an external physician panel and were determined to have non-infectious etiologies (i.e., not sepsis, and thus classified as SIRS under the definition used here). The adjudicators did not have access to the SeptiCyte RAPID test results. Aggregated clinical data for these five patients are provided in Supplement S1. However, a data use agreement did not permit sharing of individual-level data for the Ghanian participants, so their clinical values are not included in Table 1. Their SeptiScores, however, were available individually and are included in all figures.

### 2.2. SeptiCyte RAPID

SeptiCyte RAPID is a cartridge-based reverse transcription – quantitative polymerase chain reaction (RT-qPCR) test that quantifies the relative amounts of two mRNA transripts (PLAC8, PLA2G7) from human blood samples. The test has regulatory clearances for use in adults in the USA, Europe and Australia. The test was validated and shown to display consistent performance in a heterogeneous patient population of 419 ICU patients from the Netherlands and U.S.A. (Balk et al., 2024a, 2024b) [29, 30]. The validation cohort included immunosuppressed patients, and patients with low white cell counts. SeptiCyte RAPID is run on the Idylla hardware platform, using version 1.2 of the test specific software (Biocartis NV, Mechelen, Belgium). The test is performed by pipetting 0.9 mL of PAXgene-stabilized blood (corresponding to 0.24 mL of drawn blood), or 0.24 mL of EDTA anti-coagulated whole blood, into a custom cartridge which performs all assay steps including sample extraction/purification and RT-qPCR for the detection and relative quantification of the PLAC8 and PLA2G7 mRNA transcripts. The SeptiCyte RAPID test results are calculated and presented automatically through a software-generated report, which includes a quantitative score (SeptiScores, range 0–15, see **Figure 2**). The SeptiScore is calculated as the difference between the RT-qPCR Cq values for PLA2G7 and PLAC8, and is positively correlated with sepsis probability. The test has a hands-on time of ∼2 minutes and a turnaround time of ∼1 hour. In Supplement S13 we indicate the various combinations of Idylla console software version, Sep-tiCyte RAPID test software version, and SeptiCyte RAPID cartridge lots that were used in this study. The aggregate rate of invalid and failed runs (cases for which no results were generated for either PLAC8, PLA2G7, or the internal control) conformed to the test design specifications (<5% of runs).

**Figure 2.**
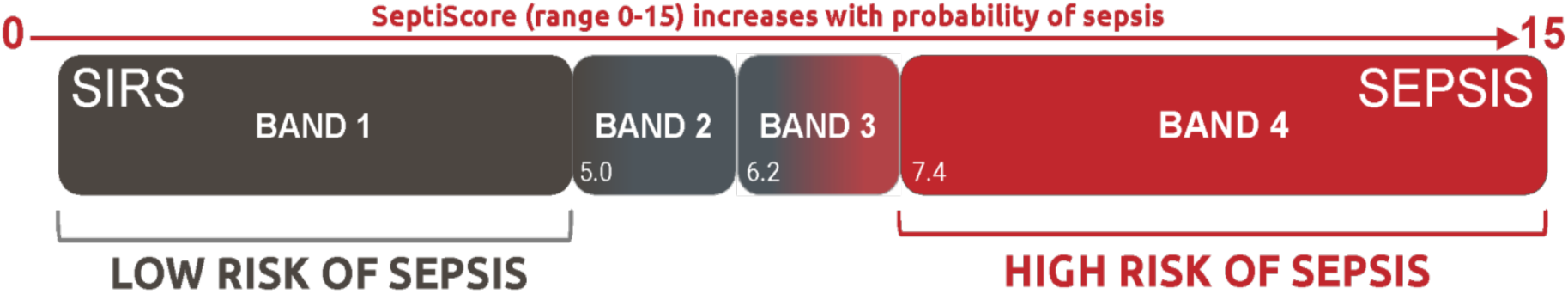
SeptiCyte RAPID Reporting. SeptiCyte RAPID provides a SeptiScore ranging from 0 to 15 and falling into one of four “SeptiScore Bands” of increasing sepsis probability. Figure adapted from Balk, et al. (2024a) [29].

### 2.3. Statistical analysis

As this is a post-hoc retrospective analysis of data from three independent studies, it has not been possible to define a ‘primary endpoint’. Instead, we test the hypothesis that a significant correlation exists between elevated SeptiScores and positive blood culture results, and between low SeptiScores and negative blood culture results.

#### 2.3.1. Conventional Statistical Tests

Conventional statistical tests were mainly conducted with the R ‘stats’ package [45]. For simple two-group comparisons, p-values were calculated using a proportions test, and for multiple-group comparisons, p-values were calculatedwith Pearson’s chi-squared test (Table 1). Point bi-serial correlations were calculated using the R package ‘ltm’ [46].

#### 2.3.2. Receiver Operating Characteristic (ROC) Curve Analysis

Receiver Operating Characteristic (ROC) curve analysis with calculation of area under curve (AUC) values [47] was performed using the pROC package in R [48]. Confidence intervals for AUC were calculated by method of DeLong et al. [49] as implemented in pROC [43]. A conventional interpretation of ROC curves was used, in which AUC 0.70-0.80 was considered acceptable, 0.80-0.90 was considered good, and AUC 0.90-1.00 was considered excellent (see e.g. Table 4 of Elkahwagy & Kiriacos, 2024) [50].

#### 2.3.3 SeptiScore Binary Cutoffs

Figure 2 presents the accepted 4-band framework for interpreting SeptiCyte RAPID test results. We also performed binary analyses, in which a single cutoff at a predefined SeptiScore boundary (5.0, 6.2, 7.4, 11.4) is used to generate 2x2 tables and sensitivity / specificity tables (Figure 5). This is most useful for understanding the behavior of the SeptiCyte RAPID test at extreme values of the SeptiScore, such as in a high sensitivity regime with binary cutoff = 5.0, or a high specificity regime with binary cutoff = 11.4. In the binary analyses, the 95% confidence intervals (CI) for sensitivity and specificity were estimated by bootstrapping [Carpenter & Bithell, 2000] [51] A two-sample proportion test was used to determine if differences between pairs of sensitivity values or specificity values were significant at the p<0.05 level.

#### 2.3.4. Probability Ratio Analysis

This study was conducted under the hypothesis that a significant correlation exists between elevated SeptiScores and positive blood culture results, and between low SeptiScores and negative blood culture results. A prediction that follows from this hypothesis is that the ratio of probabilities of BC(+) sepsis / BC(-) sepsis should increase in a monotonic fashion with increasing SeptiScore. This prediction was tested quantitatively, in the following way.

From the pooled data, the numbers of BC(+) sepsis, BC(-) sepsis, and SIRS were counted in eleven adjacent SeptiScore intervals: 0-4, 4.1-5, 5.1-6, 6.1-7, 7.1-8, 8.1-9, 9.1-10, 10.1-11, 11.1-12, 12.1-13, 13.1-15. In each interval the probabilities of BC(+) sepsis and BC(-) sepsis were then calculated as follows:

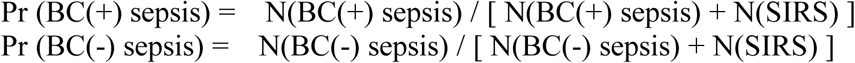

The ratio R of these two quantities was computed and plotted as a function of SeptiScore. A monotonic increase in this ratio is indicated by Ri+1 / Ri > 1 where i is the index variable for the intervals defined above. To test if a trend of monotonic increase in Ri+1 / Ri was statistically significant, the Jonckheere-Terpstra Test [46] was used, with a web applet available at www.metricgate.com

## 3. Results

### 3.1. Patient Demographic, Clinical and Laboratory Characteristics

**Table 1** presents the demographic and clinical characteristics of the patients analyzed in the present study. As described in the flow diagram of Figure 1, a total of 652 patients suspected of sepsis were originally considered, comprising 419, 167 and 61 patients from the 510k, UGANDA and SeptAsTERS studies respectively. An extra 5 patients from a Ghanaian cohort were added, to supplement the number of available SIRS patients from the UGANDA study. For 41/419 (9.8%) of patients from the 510k study and 30/167 (18.0%) of patients from the UGANDA study, no unambiguous call of sepsis or SIRS could be made; these patients were therefore deemed “indeterminate” and excluded from further analysis. Of the 322 patients deemed septic, 73 were excluded because blood cultures were either not taken or blood culture results were not recorded (73/322 = 22.7%). A total of 504 patients ultimately were analyzed in the present study (red boxes in Figure 1), comprising 247 with sepsis (consensus or site adjudicated) and available blood culture results, and 257 with SIRS (consensus or adjudicated).

Besides the geographic differences in origin between the Ugandan, 510k and SeptAsTERS cohorts, there were slight but statistically significant imbalances in the male/female ratio in both the 510k and SeptAsTERS cohorts (p < 0.04 each). A statistically significant difference (p < 0.001) was observed between the three studies for the sequential organ failure assessment (SOFA) score. Patients from the Uganda study had a lower median SOFA score (median 2) vs. patients from the 510k study (median 5) or the SeptAsTERS study (median 5).

Patient laboratory characteristics, based on sepsis / SIRS calls and blood culture status, for the three studies are presented in Supplements S2-S5. The median C-reactive protein (CRP) level was elevated in all three studies and did not differentiate sepsis from SIRS patients.

CRP was highest in the SeptAsTERS study which consisted exclusively of post-abdominal surgery patients. White blood cell (WBC) counts varied widely, from 300 to 57,000 cells / uL (510k study), 2,000 to 75,000 cells/uL (UGANDA study), and 1,300 to 63,000 cells/uL (SeptAsTERS study). In each study, the highest WBC counts were observed in the sepsis group, as opposed to the SIRS group. Median SeptiScores for patients with positive blood cultures fell in SeptiScore Band 4 (>7.4) for all three studies.

Across all three studies, a total of 85 septic patients had positive blood cultures with similar proportions of Gram-positive and Gram-negative bacterial isolates. The 510k study had six patients with mixed culture results, in which both Gram-positive and Gram-negative bacteria were isolated. A detailed breakdown of how the positive blood culture sepsis calls distribute across the SeptiScore bands, for each of the study cohorts, is presented in **Table 2**.

**Table 2:** Distribution of BC(+) sepsis calls across the Cohorts and SeptiScore Bands.

| SeptiScore Band | BC(+) sepsis |  |  |  |
| --- | --- | --- | --- | --- |
|  | 510k | SeptAsTERS | Uganda + Ghana | All cohorts |
| **Band 4** | 34 (70.8%) | 12 (80.0%) | 20 (90.9%) | 66 (77.7%) |
| **Band 3** | 9 (18.8%) | 1 (6.7%) | 1 (4.6%) | 11 (12.9%) |
| **Band 2** | 5 (10.4%) | 2 (13.3%) | 1 (4.6%) | 8 (9.4%) |
| **Band 1** | 0 | 0 | 0 | 0 |
| **BC(+) sepsis / all sepsis** | 48/305 (15.7%) | 15/59 (25.4%) | 22/142 (15.5%) | 85/506 (16.8%) |
| **Gram stain: (+)/(-)/mixed** | 20/22/6* | 6/9/0** | 11/11/0*** | 37/42/6 |

\* one Gram(+) was fungal, and one mixed Gram(+)/(-) was bacterial + fungal
\*\* two Gram(+) were fungal
\*\*\* one Gram(+) was fungal

### 3.2. Performance of SeptiCyte RAPID Compared to Blood Culture

We stratified patients on the basis of sepsis / SIRS status, blood culture result, and SeptiScores (Figure 3). In panels A, B, C of this figure, the four SeptiScore bands are separated by vertical black lines where Band 1 indicates lowest risk of sepsis and Band 4 indicates highest risk of sepsis. Panel A (red points) shows the distribution of SeptiScores (scale of 0-15) for 85 blood culture positive patients diagnosed with sepsis. An important feature to note are that most (66/85 = 78%) of the patients fell in SeptiScore Band 4, with some patients displaying very high SeptiScores. Further detail on the microbiology results for the 5 patients with highest SeptiScores (12.9-15.0) is given in Supplement S6.

**Figure 3.**
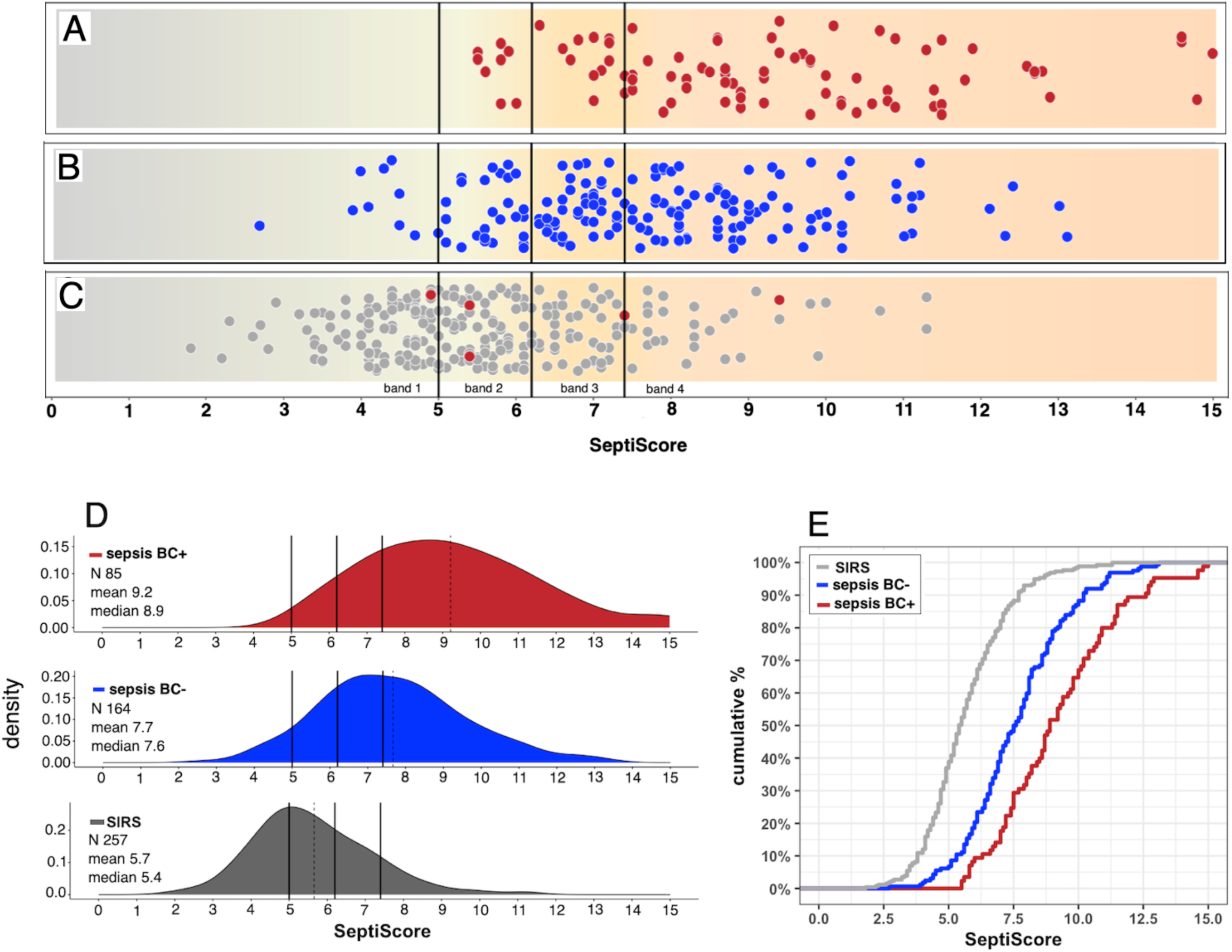
Distribution of Patients Based on RPD (sepsis / SIRS), Blood Culture Status and SeptiScore. Patients from all three cohorts are included. For A, B and C, SeptiScore (range 0-15) is shown on the horizontal axis, with the boundaries between adjacent SeptiScore bands indicated by vertical black lines at values 5.0, 6.2 and 7.4. **(A)** patients retrospectively diagnosed as sepsis with positive blood cultures (red points, n=85). **(B)** patients retrospectively diagnosed as sepsis with negative blood culture results (blue points, n=162) of which 126 (77.8%) fall into SeptiScore bands 3 and 4. **(C)** patients retrospectively diagnosed with SIRS that were blood culture negative (grey points, n=257). Also shown in this panel are three patients retrospectively diagnosed as SIRS, with positive blood cultures (red points, n=5). **(D)** Quantification of data from Panels A, B, C in terms of SeptiScore density distributions. The vertical solid black lines indicate the band boundaries at 5.0, 6.2 and 7.4. The vertical dotted lines indicate the means of the density plots. **(D)** Quantification of data from Panels A, B, C in terms of SeptiScore cumulative distributions.

A second important feature to note about Panel A is that none of the BC(+) sepsis patients fell in SeptiScore Band 1. There were, however, eight (8/85 = 9.4%) such patients with SeptiScoress in Band 2. These would be considered “false negatives” for sepsis if evaluated solely on the basis of their relatively low SeptiScores. The eight patients were spread across all three studies. Further detail on the microbiology results for these patients can be found in Supplement S7. Both Gram-negative and Gram-positive bacteria were isolated. Four of the patients (# 1, 4, 6, 8 in Table S7) received antibiotics before the blood sample was drawn for SeptiScores measurement, which might explain their low SeptiScores. Four patients (# 1, 2, 5, 6 in Table S7) displayed long blood culture times to positivity (3d, 4d, 9d, 10d) arguing for contamination; these might be better placed in the sepsis BC(-) category of Panel B (below).

However, patient #3 (*S. anginosus* from central catheter, time to positivity 1 day) and Patient # 7 (*K. pneumoniae*, time to positivity 41 hours) were not obvious contaminants, were not pre-treated with antibiotics, and fell below the 10^th^ percentile of the SeptiScore cumulative distribution for BC(+) sepsis (see Fig 3D below), and therefore should be flagged for further investigation.

Panel B (blue points) shows the distribution of SeptiScores for 162 sepsis patients that had blood cultures ordered with negative results, or that were positive for cultures other than from blood. Of the 162 patients retrospectively diagnosed with sepsis and with negative blood cultures, 126 (77.8%) had SeptiScores in Bands 3 or 4. Some of the BC(-) sepsis patients had quite extreme Band 4 SeptiScores; five patients with highest SeptiScores (12.1-13.1) are detailed in Supplement S8.

Another point to note is that 10 BC(-) sepsis patients had Band 1 SeptiScores. These comprise 6.2% of the 162 patients in the BC(-) sepsis category. Further detail on these patients can be found in Supplement S9. Patients of the types represented in this table would seem to require further detailed investigation, because if they were truly septic, they would have been missed by an alert system based on either blood culture or SeptiCyte RAPID.

Panel C shows the distribution of SeptiScores for SIRS patients that were blood culture negative (grey dots) and for five blood culture positive SIRS patients (red points). Of note is the high proportion of SIRS patients in Bands 1 and 2 (n=175/ 263 = 66.5%). Further detail on the microbiology results for the five BC(+) SIRS patients can be found in Supplement S10. The bacteria isolated from the three BC(+) SIRS patients with lowest SeptiScores were considered by the RPD panelists to be not clinically relevant and possibly contaminants: *Bacillus spp.* (SeptiScore 4.9); *Fusobacterium* (8 days to positivity, no antibiotics given, SeptiScores 5.4); *coagulase negative Staphylococcus aureus* (6 days to positivity, no antibiotics given, SeptiScore 5.4).

There were also some BC(-) patients deemed SIRS, but with Band 4 SeptiScores. These are represented by the rightmost grey points in Panel C. Further detail on a selection of these patients is provided in Supplement 11.

Panels D and E present a quantification of the data from Panels A, B, C in terms of SeptiScores density distributions and cumulative distributions, respectively. There is clearly an offset of the distributions in the direction SIRS < sepsis BC(-) < sepsis BC(+).

Based on the data presented in Figure 3, we next evaluated the diagnostic performance of SeptiCyte RAPID for differentiating patients that were retrospectively diagnosed septic and blood culture positive, from those retrospectively diagnosed with SIRS in a patient population initially suspected of sepsis. **Table 3** summarizes the results of this evaluation, according to SeptiScore Band. Of the 348 patients considered in this table, 85 were both septic and blood culture positive, and 263 were determined to have SIRS. Patients with a SeptiScore falling between 7.4 and 15 (Band 4) had a positive likelihood ratio (LR+) of 5.88 for blood culture positive sepsis (as compared to SIRS). In contrast, patients with a SeptiScore falling between 0 and 4.9 (Band 1) had an LR+ of 0. Further detail, including LR+ calculations, are provided in Supplement S12. A more complete analysis of multi-cohort data in terms of positive and negative likelihood ratios will be presented in a separate publication.

**Table 3.** Diagnostic performance of SeptiCyte RAPID, for BC(+) sepsis versus SIRS, in terms of Positive Likelihood Ratio (LR+). For this table calculations were based on 85 patients that were blood culture positive and diagnosed with sepsis, and 263 patients that were diagnosed with SIRS (including 3 SIRS patients with positive blood cultures). Patients with a SeptiScore falling between 7.4 and 15 (Band 4) had an LR+ of 5.88 for blood culture positive sepsis relative to SIRS. Patients with a SeptiScore falling between 0 and 4.9 (Band 1) had an LR+ of 0 for blood culture positive sepsis relative to SIRS.

| SeptiCyt<sup>®</sup> RAPID Band | Sepsis and BC(+) | SIRS | Total Patients | LR+ |
| --- | --- | --- | --- | --- |
| Band 4 (7.4 – 15) | 66 | 34 | 100 | 5.88 |
| Band 3 (6.2 – 7.3) | 11 | 50 | 61 | 0.66 |
| Band 2 (5.0 – 6.1) | 8 | 78 | 86 | 0.31 |
| Band 1 (0 – 4.9) | 0 | 95 | 95 | 0 |
| **Total** | **85** | **257** | **348** |  |

**Table 4:**
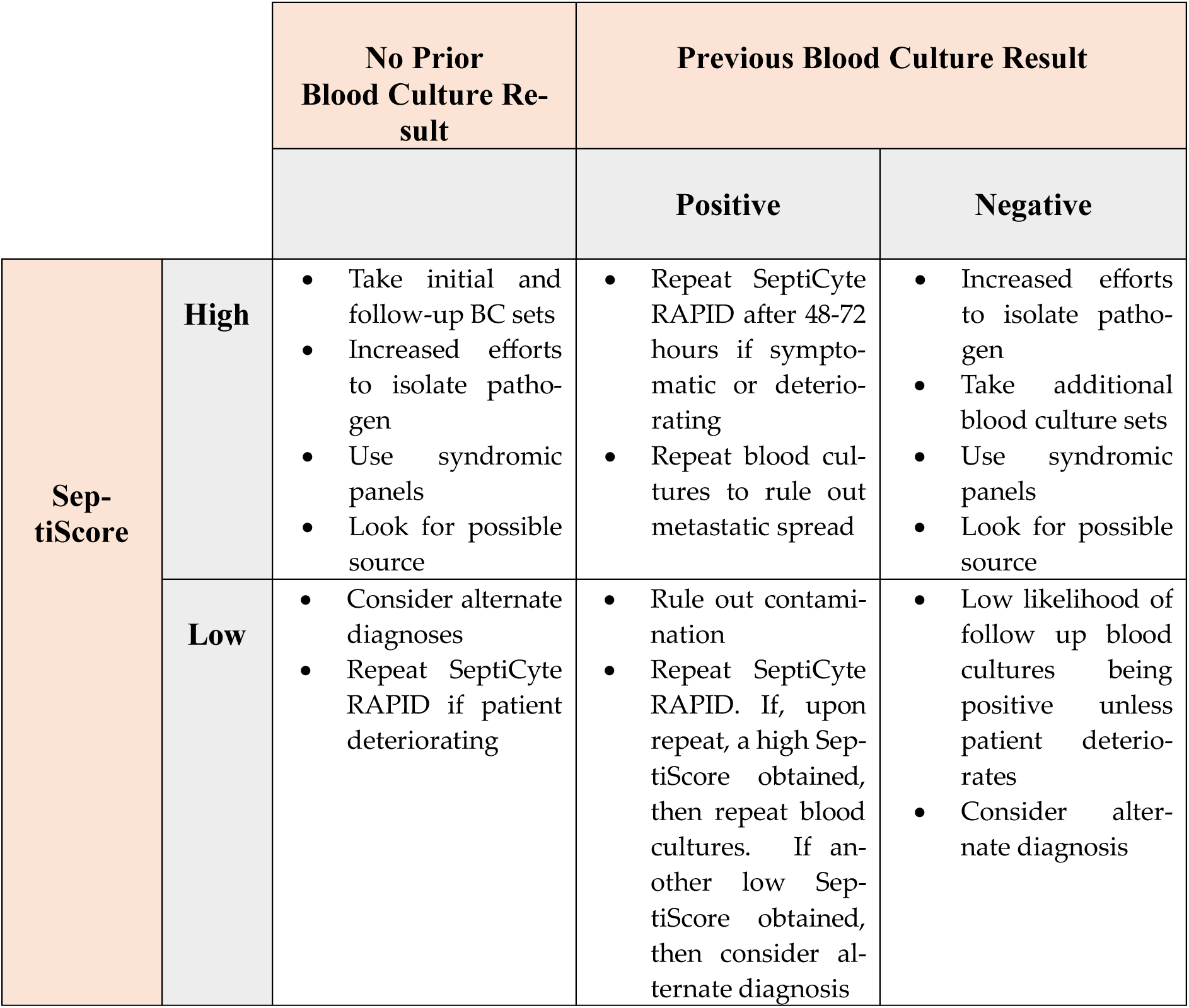
Potential Clinical Uses of SeptiCyte RAPID for Blood Culture Practice.

|  |  | No Prior Blood Culture Result | Previous Blood Culture Result |  |
| --- | --- | --- | --- | --- |
|  |  |  | Positive | Negative |
| SeptiScore | High | Take initial and follow-up BC sets<br>Increased efforts to isolate pathogen<br>Use syndromic panels<br>Look for possible source | Repeat SeptiCyte RAPID after 48-72 hours if symptomatic or deteriorating<br>Repeat blood cultures to rule out metastatic spread | Increased efforts to isolate pathogen<br>Take additional blood culture sets<br>Use syndromic panels<br>Look for possible source |
|  | Low | Consider alternate diagnoses<br>Repeat SeptiCyte RAPID if patient deteriorating | Rule out contamination<br>Repeat SeptiCyte RAPID. If, upon repeat, a high SeptiScore obtained, then repeat blood cultures. If another low SeptiScore obtained, then consider alternate diagnosis | Low likelihood of follow up blood cultures being positive unless patient deteriorates<br>Consider alternate diagnosis |

Note that the LR+ values for BC(+) sepsis depend upon the composition of the group to which BC(+) sepsis is being compared. In our analysis, the comparator is the BC(-) SIRS group [patients of Fig 3c], consistent with the Intended Use of the SeptiCyte RAPID test (discrimination of sepsis vs. SIRS). Understandably, the LR+ values for BC(+) sepsis would be decreased if the BC(-) comparator group were expanded to include BC(-) sepsis patients [i.e. Fig 3b patients mixed in with Fig 3c patients]. However, in practice this would not be a meaningful comparison, because clinicians would be expected to use SeptiCyte to discriminate between sepsis and SIRS in accordance with the test’s Intended Use, not to discriminate between BC(+) sepsis and BC(-) sepsis.

We next performed additional analyses of SeptiCyte RAPID performance, including calculations of area under the receiver operating characteristic curve (ROC AUC), sensitivity, specificity and Youden’s index, using data from the combined three study cohorts. Figure 4A reproduces the data of Figure 2 in the form of box-whisker plots. Figure 4B presents three ROC curves for SeptiCyte RAPID performance, for sepsis vs. SIRS discrimination, depending on the septic patients’s blood culture results. The red curve represents BC(+) sepsis versus SIRS AUC = 0.91). The blue curve represents BC(-) sepsis versus SIRS (AUC = 0.79).

**Figure 4.**
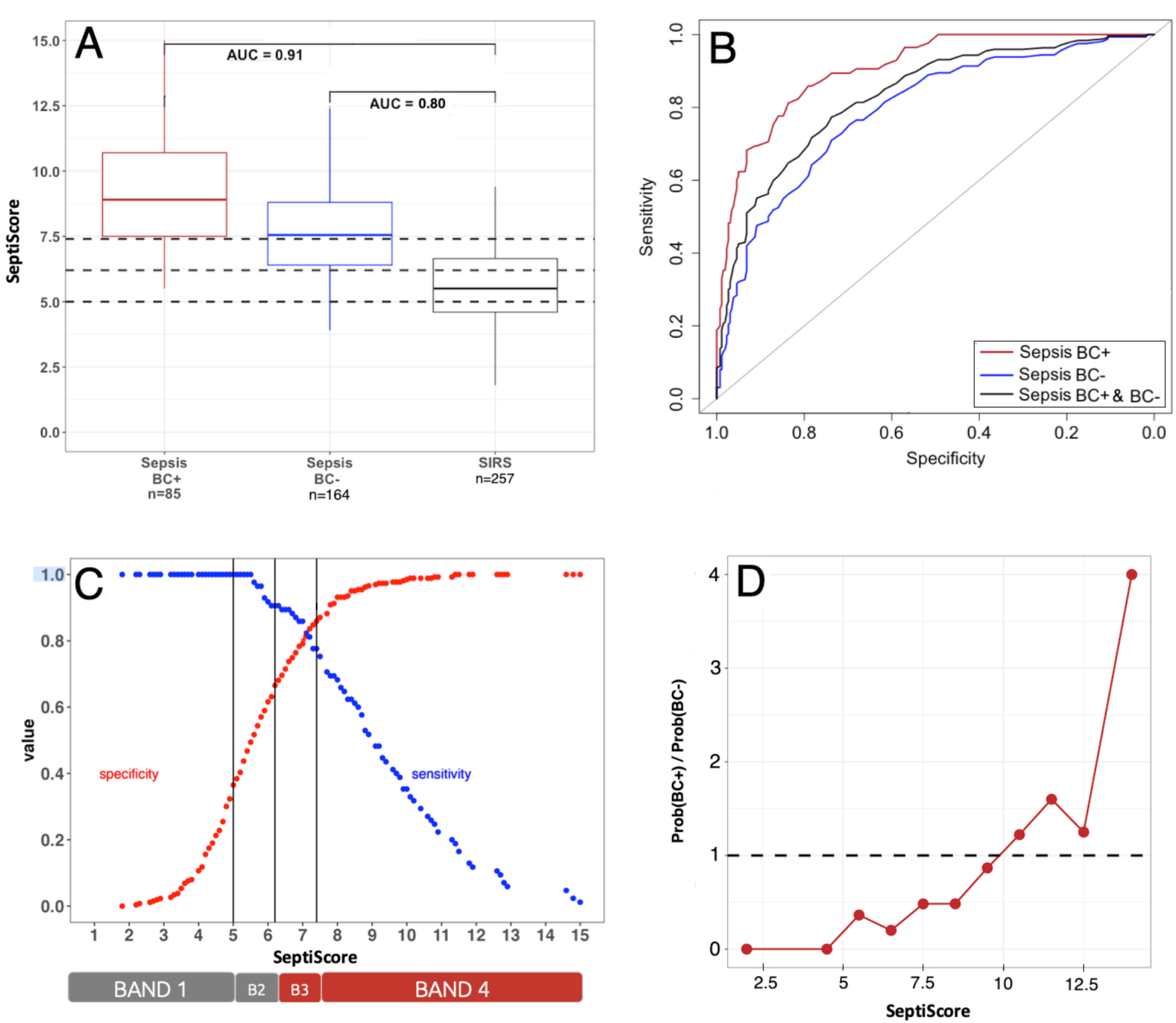
SeptiCyte Performance in Blood Culture Positive Sepsis Patients. **(A)** Box and whisker plot showing performance of SeptiCyte RAPID when comparing sepsis patients that were either BC(+) (85 by consensus) or BC(-) (164 by consensus) to 257 SIRS patients. Data compiled from all three studies. From ROC analysis (Panel B) the calculated AUC for BC(+) sepsis vs. SIRS was 0.91 and the calculated AUC for BC(-) sepsis vs. SIRS was 0.80. **(B)** SeptiCyte RAPID ROC curves for BC(+) sepsis vs. SIRS (red, AUC 0.91, 95% CI 0.87-0.94), for BC(-) sepsis vs. SIRS (blue, AUC = 0.80, 95% CI: 0.75-0.84), and for pooled sepsis (irrespective of blood culture results) vs. SIRS (black, AUC = 0.84, 95% CI: 0.80-0.87).**(C)** Binarized model in which sensitivity (blue) and specificity (red) are plotted as a function of cutoff SeptiScore for dataset consisting of BC(+) sepsis (N=85) vs. SIRS (N=257). Binary cutoffs at 5.0, 6.2, 7.4 are shown as vertical black lines. **(D)** The ratio of probabilities for BC(+) sepsis versus BC(-) sepsis, as a function of SeptiScore. The data points in this panel are calculated from the numbers of BC(+) sepsis, BC(-) sepsis, and SIRS in the following eleven SeptiScore intervals: 0-4, 4.1-5, 5.1-6, 6.1-7, 7.1-8, 8.1-9, 9.1-10, 10.1-11, 11.1-12,12.1-13, 13.1-15, and are plotted at the interval midpoints. For septic patients, increasing SeptiScore correlates positively with an increase in relative probability of being BC(+) as opposed to BC(-). The dotted line indicates a probability ratio of 1.0. For SeptiScores above 10 the probability of a septic patient being being BC(+) is higher than that of being BC(-). The correlation between increasing SeptiScore and increasing probability ratio is statistically significant according to the Jonckheere-Terpstra test statistic value J = 52.5 and p-value: 9.92 x 10^-5^.

**Figure 5.**
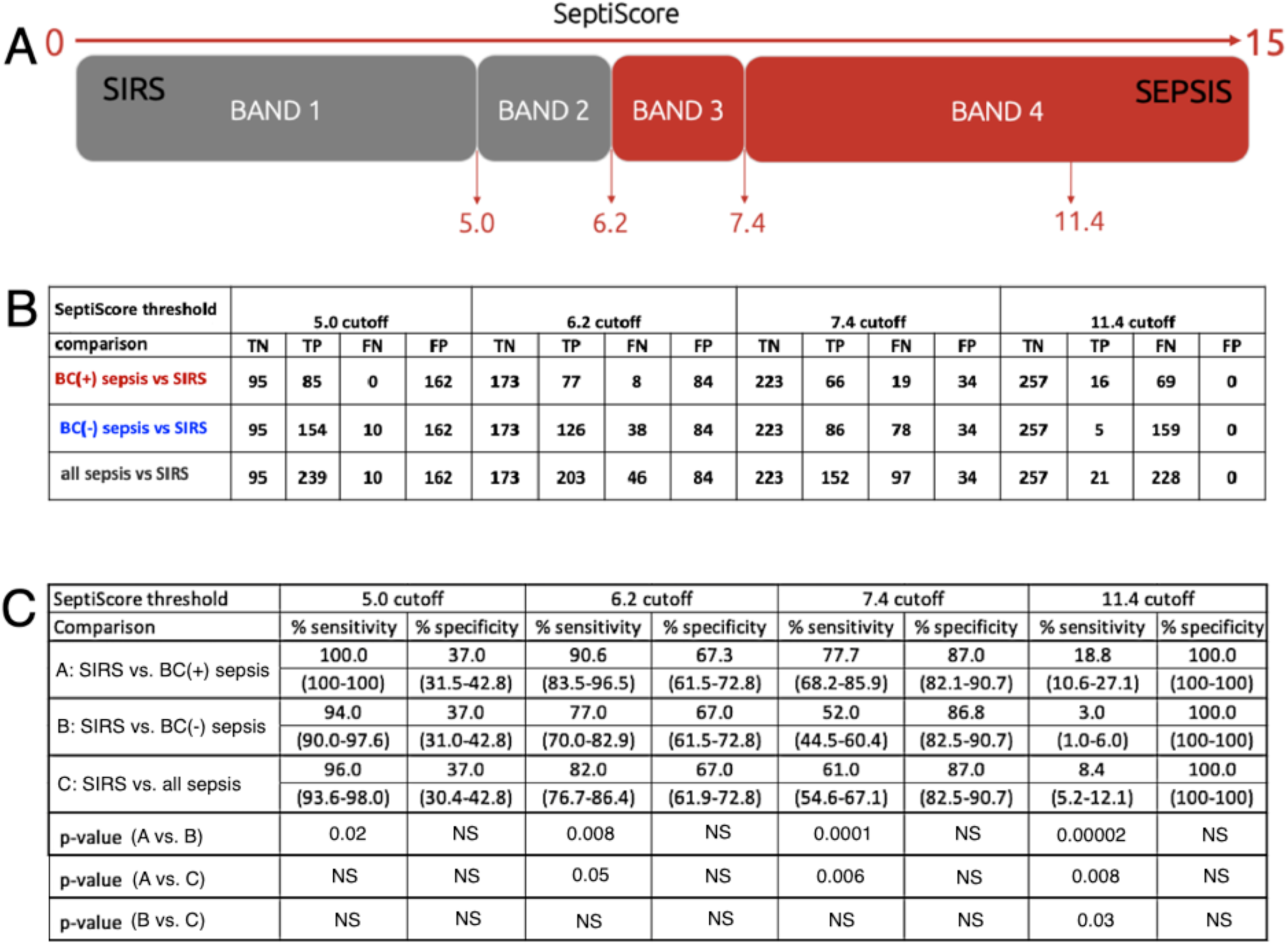
Sensitivity and Specificity of SeptiCyte RAPID for Sepsis BC(+) or Sepsis BC(-) Versus SIRS. This figure presents binarized classification models, with cutoffs at the predefined SeptiScore boundaries (5.0, 6.2, 7.4, 11.4). (A) Schematic diagram. (B) Numbers of true negatives (TN), true positives (TP), false negatives (FN) and false positives (FP) for each binary cutoff value. (C) Calculated sensitivity and specificity values for BC(+) sepsis and BC(-) sepsis vs. SIRS for each SeptiCyte RAPID band boundary, and for a cutoff at 11.4 where SeptiCyte RAPID has 100% specificity (no patients called SIRS below this SeptiScore). The 95% confidence intervals (shown in parentheses) were calculated by bootstrapping and the p-values were calculated with a two-sample proportions test, as described in Materials and Methods. Abbreviation: NS, not significant (p > 0.05).

Finally the black curve represents sepsis irrespective of blood culture results, versus SIRS (AUC = 0.83). Figure 4C shows sensitivity (blue) and specificity (red) curves as a function of the SeptiScore, for the BC(+) versus SIRS patients. The two curves cross at SeptiScore of 7.0 (Youden’s index). Band boundaries at 5.0, 6.2, 7.4 are shown as vertical black lines. **Figure 4D** presents a plot of 11 incremental calculations of the relative probabilities of BC(+) sepsis / BC(-) sepsis, as a function of SeptiScore. As the SeptiScore increases, the relative probabilty of a septic patient being BC(+) as opposed to BC(-) also increases. The correlation between increasing SeptiScore and increasing probability ratio is statistically significant according to the Jonckheere-Terpstra trend test [52] as indicated by test statistic value J = 52.5 and p-value: 9.92 x 10^-5^. At a SeptiScore of 10, the ratio of probabilities increases above 1 (dotted line in figure). That is, septic patients with SeptiScores >10 have a higher probability of being BC(+) compared to BC(-).

Figure 5 presents sensitivity and specificity calculations for BC(+) sepsis and BC(-) sepsis versus SIRS, for binarized models with single cutoffs at the predefined SeptiScore Band boundary (5.0, 6.2, 7.4) and also at 11.4. This figure is derived from the more complete ROC curve analysis of Figure 4B. For a binary cutoff at SeptiScore 5.0 the sensitivity was 100%, and for a binary cutoff at SeptiScore 11.4 the specificity was 100%. That is, based on the patients considered in this study, no patients with SeptiScores less than 5.0 were retrospectively BC(+) sepsis, and no patients with SeptiScores greater than 11.4 were retrospectively SIRS.

## 4. Discussion

The current reference standard for diagnosing BSI is blood culture. However, blood culture practice and interpretation of results have a number of limitations including low positive rates, contamination and a lack of timeliness. Downstream implications from these limitations include unnecessary antibiotic use, an increase in overall mortality and antimicrobial resistance, increased pharmacy and laboratory costs, and length of hospital stays (Butler 2023, Fabre 2025, Dempsey 2019) [13, 18, 20]. A number of interventions as part of blood culture diagnostic stewardship have been described including careful patient selection and efforts to keep contamination rates less than or equal to a target rate of 1% of all blood cultures taken (Fabre 2020, Schinkel 2023, Doern 2019, CLSI M47) [53–56]. Therefore, it can be argued that blood culture practice is currently suboptimal and there is a need for improved BC stewardship (Fabre, 2025) [18].

In this study, we conducted a post-hoc retrospective analysis of patients suspected of sepsis, representing diverse populations across multiple continents (North America, Europe, Africa). We have shown a positive correlation between sepsis likelihood, measured using SeptiCyte RAPID, and likelihood of blood culture positivity. From our study of 504 patients, of which 16.9% (n=85) were blood culture positive, our results showed that patients with SeptiScores greater than 10 had a higher probability of being BC(+) sepsis than BC(-) sepsis. Further, patients with SeptiScores less than 5.0 had a 0% probability of being BC(+) sepsis (Figure 4C). On the basis of this finding, we hypothesize that positive blood cultures in patients with SeptiScores less than 5.0 are likely to be either contaminants or not relevant to a sepsis diagnosis.

The blood culture positivity rate in our study (16.9%) was higher than the 5.6% to 7.3% recently reported in a large study involving 362,327 blood cultures conducted between 2019 and 2020 and in 19 USA hospitals (Fabre 2025) [18]. The higher positivity rate in our study may relate to unintentional bias in patient selection, since the patients in all three cohorts examined here were enrolled on the basis of being critically ill. It might also reflect an unduly high rate of culture contamination. (For comparison, in the 2025 Fabre study [18] the contamination rate was 1.5% of all blood cultures taken which equates to ∼24% contamination rate for positive cultures.) The overall contamination rate in our study could not be accurately calculated because a blood culture was not taken from every patient. However, out of a total of 85 blood culture positive results there were 3 retrospectively diagnosed with SIRS and with low SeptiScores and considered to be either contaminants or not relevant to a sepsis diagnosis (3.5% of all positive cultures).

As shown in Figures 4A and 4B, the ROC AUC for using SeptiScore to distinguish SIRS from BC(+) sepsis was 0.91. The corresponding likelihood ratio (LR) for blood culture positivity among patients with Band 4 SeptiScores was 5.88. Notably, no BC(+) patients retrospectively diagnosed with sepsis had SeptiScores in Band 1. Collectively, these findings indicate that SeptiCyte RAPID demonstrates strong performance for discriminating BC(+) sepsis from SIRS. Although its ability to discriminate BC(–) sepsis from SIRS was lower, it remained diagnostically meaningful (ROC AUC 0.80). In our study, all patients with SeptiScoress >11.4 were either BC(+) septic or BC(–) septic. Further inspection of the data revealed four patient subgroups warranting additional consideration.

1. <u>BC(-) sepsis with high SeptiScores:</u> Of the 164 patients classified as septic by RPD with negative blood cultures, 85/164 (51.8%) had SeptiScores in Band 4, and 126/164 (78.6%) had SeptiScores in either Band 3 or Band 4 (Figure 3B). Thus SeptiScores tended to be elevated even in cases of apparent non-bacteremic sepsis. Cases of culture-negative sepsis are well known and have received significant attention [Phua et al., 2013; Thorndike & Kollef, 2020; Li et al., 2021; Chang et al, 2024]. [57–60]. It is recognized that some septic events may not be bacteremic in nature, and may instead be due to localized (non-bloodborne) bacterial infections, or to infections from other types of pathogens (viral, fungal, parasitic) [Chang et al. 2024; Lee e al. 2024; Xu et al., 2024] [60–62]. Given these possibilities high SeptiScores could potentially help guide diagnostic stewardship by providing motivation for more aggressive attempts to identify causative pathogens, including additional blood cultures, culturing of patient samples other than blood, or use of rapid molecular viral panel tests.
2. <u>BC(+) sepsis with low SeptiScores:</u> Of the 85 patients classified as septic by RPD with positive blood cultures, none had SeptiScores in Band 1, and eight (8/85 = 9.4%) had SeptiScoress in Band 2 (Figure 3A). SeptiScores in Band 2 would be considered relatively low values for patients who were truly septic and blood culture positive. However, closer examination of the microbiology results for these eight patients showed that four had prior antibiotic exposure, which may have influenced the patient systemic immune response to bacterial threat. Additionally, four of the patients had long time-to-positivity (3, 4, 9 and 10 days), which may indicate that the positive blood culture results were in fact due to contamination. Such results suggest that many factors need to be taken into consideration when interpreting the clinical relevance of SeptiCyte RAPID and blood culture results, including the timing of blood draws, effect of antibiotics, time to positivity, organism isolated, potential for contamination, other clinical parameters and laboratory results, and patient trajectory (whether the patient is getting better or worse).
3. <u>BC(-) SIRS with high SeptiScores:</u> Of the 252 patients classified as SIRS by RPD with negative blood cultures, thirty two exhibited Band 4 SeptiScores (32/252 = 12.7%) (Figure 3C). This suggests that, in some cases, factors other than sepsis may lead to elevated SeptiScoress, consistent with the view that SeptiScores should not be interpreted in isolation, but preferably together with other available clinical data. By way of example (see S11 for further detail), a patient with a high (Band 4) SeptiScore of 7.7 with no initial confirmed site of infection nonetheless had an RPD call of SIRS. The patient was prescribed a combination of antibiotics for 2.6 days after presentation, but ultimately (at 2.6 days) was diagnosed by PCR as having Epstein-Barr Virus in blood. A high SeptiScore in this patient, especially if obtained early in a diagnostic workup, may have provided diagnostic stewardship with respect to determining a viral etiology.
4. <u>BC(+) SIRS with low SeptiScores:</u> Of the 257 patients classified as SIRS by RPD, three had SeptiScores in Band 1 or Band 2 and had positive blood cultures (3/257 = 1.2%) (Figure 3C). By way of example (see S10 for further detail), one patient with a low (Band 2) SeptiScores of 5.4 had a positive blood culture for *Staphylococcus aureus*. Normally, identification of this known pathogen from blood culture might suggest that the patient had sepsis of a bacterial etiology. However, no site of infection was identified initially, there was a long time-to-positivity (6 days), and no antibiotics were prescribed. The patient was ultimately called SIRS by RPD, and the low SeptiScore was consistent with this diagnosis. Contamination would be most likely explanation for blood culture positivity in this case.

A practical point to note is that SeptiCyte RAPID results are available within one hour and in the same time frame as WBC data. Such a timeframe is significantly shorter than reported median times to blood culture positivity of 15.7 hours (Lambregts et al., 2019) [15].

Furthermore, SeptiCyte RAPID results are available at least 12 hours prior to gaining any blood culture results and within the 3-hour period recommended by the Surviving Sepsis Campaign (Evans et al., 2021) [63] for administration of antibiotics for patients suspected of sepsis (without shock).

Some potential clinical uses of SeptiCyte RAPID for informing blood culture practice are presented in **Table 4**. However, we recognize that clinical utility of SeptiCyte RAPID for informing blood culture practice still needs to be rigorously demonstrated in a prospective, cluster-randomized stewardship trial, to include safety endpoints such as missed bacteremia, ICU transfer, and 7/30-day mortality.

For patients with high SeptiScores (>7.4, Band 4) and positive blood culture results, SeptiCyte RAPID could potentially be repeated to confirm a diagnosis and to rule out metastatic spread. For patients with high SeptiScores and negative blood culture results, more intense efforts could be made to identify a causative pathogen to include the use of multiple blood culture sets and molecular diagnostic syndromic panels. High SeptiScores might also be an indication the antibiotic treatment is not effective.

For patients with low SeptiScores (<5.0, Band 1) and positive blood cultures, contamination could be considered, or alternatively bacteremia in the absence of sepsis. Examples of transient or even persistent bacteremia in the absence of sepsis are well known, e.g. due to periodontitis ore periodontal procedures (Everett & Hirschmann, 1977; Takai et al., 2005; Lockhart et al., 2008; Horliana et al., 2014) [64–67] For patients with low SeptiScores and negative blood culture results, alternative diagnoses (other than sepsis) should be considered.

Further clinical utility for SeptiCyte RAPID could lie in determining whether follow-up blood cultures (FUBC) are needed. It is likely that, in standard clinical practice, additional blood cultures would be drawn if a patient continued to deteriorate. FUBC are generally also collected after the occurrence of a positive blood culture with a known pathogen that matches the clinical presentation, to monitor the persistence or clearance of bacteraemia, or to determine antibiotic sensitivity or resistance profiles. FUBCs are suggested in the management of *Staphylococcus aureus* bacteraemia, endocarditis, and candidemia (Liu 2011, Baddour 2015, Pappas 2016 [68–70]. However, there is controversy as to the utility of FUBC in patients with Gram-negative BSI (Ong et al., 2024). [71] SeptiCyte RAPID results obtained within 1-2 hours of testing could provide guidance on whether, or to what extent FUBCs are warrented.

Our study has several limitations. First, despite enrolling over 500 patients, there were only 85 septic patients with confirmed positive blood cultures (85/506 = 16.8% positivity), which may limit the generality of conclusions drawn. However, the study spanned several geographic regions (North America, Europe, Africa) and enrolled patients with diverse comorbidities, and with WBC counts ranging from neutropenic to above normal. Similar blood culture positivity rates, isolated pathogens and SeptiScore distributions for each of the three separate studies suggests that our results and conclusions could be applied more broadly. It should be recognized, though, that the African cohort was recruited and assessed with a relatively well-controlled clinical trial, which may be atypical of the usual level of care available in this LMIC setting. An in-depth comparison of results from the three geographic areas (North America, Europe, Africa) is ongoing and will be presented in a future publication. A second limitation was that in the 510k study, blood cultures were taken at the discretion of the attending clinician. That is, not every enrolled patient had a blood culture taken. Patients with complicated diagnoses such as pneumonia with possible sepsis may not always have triggered blood culture orders [72], and patients with low-level bacteremia may not have undergone extensive enough blood sampling [73], and so may have been ‘missed’. Calculations of blood culture positivity and contamination rates are therefore not necessarily accurate for a population of patients merely *suspected* of sepsis. Third, exact timing for blood culture and SeptiCyte RAPID sampling was not known for all patients. Therefore, the influence of antibiotics on blood culture and SeptiCyte RAPID results could not be established with certainty, nor could an exact correlation between blood culture and SeptiCyte RAPID results be determined. A fourth limitation was the use of different reference methods to adjudicate sepsis versus SIRS across study sites: the 510k and SeptAsTERS cohorts were assessed via the RPD method, while the Ugandan cohort relied on site-based adjudication. As a result, estimates of SeptiCyte RAPID’s performance for distinguishing BC(+) sepsis from confirmed SIRS may be less certain for the Ugandan cohort than for the 510k and SeptAsTERS cohorts. Fifth, there may have been some selection bias, for example under the decision to exclude patients with “indeterminate” diagnoses instead of forcing these patients into the sepsis or SIRS categories (Figure 1). Sixth, there is disagreement in the field about the utility of using “SIRS” as a reference category in the diagnosis of sepsis. The 2016 Sepsis-3 consensus reoriented sepsis definitions toward organ-dysfunction–based criteria, thereby reducing the prominence of SIRS in this capacity (Singer et al., 2016) [74]. Nevertheless, many clinicians, specialty societies, and investigators continue to support the use of SIRS criteria for early clinical screening, for maintaining continuity with historical epidemiologic data, or for specific care settings where SIRS retains operational value (Balk, 2014; Simpson, 2018; Schefzik et al., 2023). [75–77]. Because the field has not reached a uniform position on the role of SIRS in defining or contextualizing sepsis, some uncertainty is introduced into the quantitative comparisons presented here, and this lack of consensus may limit the degree to which the present analysis attains broad, universal acceptance. Finally, SeptiCyte RAPID provides a likelihood of sepsis based on a SeptiScore range of 0-15 and is pathogen agnostic. In future prospective usage of the test, high SeptiScores could be due to a viral or parasitic infection rather than a bacterial or fungal infection as detected by culture. Therefore, caution would need to be taken with respect to interpreting high SeptiScores which indicate higher likelihood of sepsis rather than just culture-positive bacterial or fungal infection.

## 5. Conclusions

SeptiCyte RAPID test results (SeptiScores) are positively correlated with the likelihood of blood culture positivity. This finding could facilitate diagnostic stewardship with respect to patient selection and interpretation of positive blood culture results. Early diagnosis and identification of causative pathogens in patients suspected of BSI and sepsis is expected to improve patient outcomes, through early and more targeted treatment. Significant potential benefits are expected in diverse clinical settings, including in poorly resourced or challenging environments such as LMIC healthcare settings, mass-casualty scenarios, and prolonged field care settings during military operations. Further work could involve determining the generalisability of these results, including whether SeptiScores correlate to the likelihood of different types of infection (e.g. Staphylococcus, endocarditis, Gram negative BSI, viruses, fungi, malaria). Ultimately the clinical utility of SeptiCyte RAPID for diagnostic stewardship will be established through interventional studies and measurement of impact on patient outcomes, hospital costs, and antibiotic stewardship.

## Supplementary Materials

Supplements S1-S13 have been combined into a single pdf file, which is available for download.

## Author Contributions

Conceptualization, R.B.B., T.D.Y., K.N.; methodology, A.W., K.N.; software, K.N; validation, A.W., M.D., M. vdF., K.N.; formal analysis, A.W., M.D., M.vdF., K.N., T.D.Y., R.B.B.; investigation, A.W., M.G., S.A., M.D., M.vdF., K.N., R.B.B.; resources, D.C., M.D., M.vdF., R.B.B.; data curation, K.N.; writing—original draft preparation, R.B.B., T.D.Y.; writing—review and editing, A.W., M.G., D.C., H.K., S.O., S.A., A.Wa. P.W., F.K., G.O., N.A., M.D., M.vdF., M.J.S., J.A.G., N.R.A., N]K.N., S.C., T.D.Y., R.B.B.; visualization, K.N., T.D.Y.; supervision, D.C., R.B.B.; project administration, D.C., M.D., M.vdF., S.C., R.B.B.; funding acquisition, D.C., N.A., M.D., M.vdF., R.B.B. All authors have read and agreed to this version of the manuscript.

## Institutional Review Board Statement

All research reported in this study was conducted in accordance with the Declaration of Helsinki, and in accordance with relevant guidelines and regulations. Informed Consent was obtained from all subjects or their legal representatives. The SeptAsTERS component of this study was approved by the Ethics Committee of the Medical Faculty of Heidelberg University (S-118/2021; approval date January 4, 2021) and was registered in the German Clinical Trials Register (DRKS00024891; registration date August 4, 2021) prior to enrolment. The Uganda component of this study was approved under IRB NMRC.2016.0004-GHA and NMRC.2017.0001 (final IRB approval date March 3, 2019). The Ghanaian supplement to this study was conducted under protocol number NMRC.2016.0004-GHA (“An Observational Study of Sepsis in Kumasi, Ghana”) and approved by the Komfo Anokye Teaching Hospital IRB and the Naval Medical Research Command (NMRC) IRB prior to enrolment, in compliance with all applicable US Federal regulations governing the protection of human subjects (final IRB approval date June 14, 2017). The 510k component of this study was conducted under the following approvals. Ethics approval for the MARS trial was given by the medical ethics committee of AMC, Amsterdam (approval # 10-056C, 16 June 2010). Ethics approvals for the VENUS trial were given by the relevant Institutional Review Boards as follows: Intermountain Medical Center/Latter Day Saints Hospital (approval # 1024931, 21 January 2014); Johns Hopkins Hospital (approval # IRB00087839, 28 January 2016); Rush University Medical Center (approval # 15111104-IRB01, 11 March 2016); Loyola University Medical Center (approval # 208291, 10 March 2016); Northwell Healthcare (approval #16-02-42-03, 1 April 2016). Ethics approvals for the NEPTUNE trial were given by the relevant Institutional Review Boards as follows: Emory University (approval # IRB00115400, 3 December 2019); Grady Memorial Hospital (approval # 00-115400, 14 January 2020); Rush University Medical Center (approval # 19101603-IRB01, 16 January 2020); University of Southern California Medical Center (approval # HS-19-0884-CR001, 10 February 2020).

This meta-analysis used individual-level data from four previously IRB-approved studies. At each originating study site, datasets were de-identified prior to pooling, with both direct and indirect identifiers removed to prevent investigator access to participant identities. The original approvals allowed use of de-identified data for research “related to SeptiCyte RAPID or sepsis” (510k study); for research “limited to the field of sepsis and severe infection in intensive care medicine” (SeptAsTERS study); or for research on “identification of diagnostic or prognostic markers of sepsis” (Ugandan and Ghanaian studies). Consistent with the U.S. Common Rule (45 CFR 46), EU GDPR (Regulation 2016/679), and CIOMS (2016) International Ethical Guidelines for Health-related Research Involving Humans, secondary analysis of de-identified data does not constitute human subjects research and did not require additional ethics approval.

## Informed Consent Statement

Informed consent was obtained from all subjects involved in the study, or from their legal representatives.

## Data Availability Statement

The datasets used and/or analyzed during the current study are Available from the corresponding authors upon reasonable request.

## Data Availability

All data produced in the present study are available upon reasonable request to the authors

## Acknowledgments

We thank Ernesto Santa Ana (Austere Environments Consortium for Enhanced Sepsis Outcomes (ACESO), Henry M. Jackson Foundation for the Advancement of Military Medicine, Inc., Bethesda, MD, USA) for expert help running the SeptiCyte RAPID test. We thank Dr. Irene Hannet (Immunexpress Inc.) for critical reviews of the manuscript.

N.A is a military service member. This work was prepared as a part of official duties. Title 17 U.S.C. 105 provides that Copyright protection under this title is not available for any work of the United States Government. Title 17 U.S.C. 101 defines a U.S. Government work as a work prepared by a military service member or employee of the U.S. Government as part of a person’s official duties. The views expressed in this article reflect the results of research conducted by the authors and do not necessarily reflect the official policy or position of the Henry M. Jackson Foundation for the Advancement of Military Medicine, Inc., the Department of the Navy, Department of the Army, Department of Defense, nor the United States Government. References to non-federal entities or their products do not constitute or imply Department of Defense endorsement of any company, product or organization.

## Abbreviations

The following abbreviations are used in this manuscript:

ANOVA: Analysis of variance
AUC: area under [ROC] curve
BC: blood culture(s)
BSI: bloodstream infection
CI: confidence interval
Cq: threshold-crossing cycle (in qPCR)
CRP: C-reactive protein
ED: Emergency department
FN: False Negative
FP: False Positive
F: female
FDA: Food and Drug Administration (USA)
FUBC: follow-up blood culture
ICU: Intensive care unit
LMIC: Lower and middle income country
LR+: positive likelihood ratio
M: male
mL: milliliter
ND: not determined
NS: not significant
PLA2G7: Phospholipase A2 group VII (mRNA transcript)
PLAC8: Placenta-associated 8 (mRNA transcript)
ROC: receiver operating characteristic (curve)
RPD: Retrospective Physician Diagnosis
RT-qPCR: reverse transcription – quantitative polymerase chain reaction
SIRS: systemic inflammatory response syndrome
SOFA: sequential organ failure assessment (score)
TN: True Negative
TP: True Positive
WBC: white blood cell

