## Supplements for "Likelihood of blood culture positivity using SeptiCyte RAPID"

Supplement S1. Aggregate Characteristics of Ghanaian SIRS Patients.

Supplement S2. Laboratory Parameters for BC(+) Sepsis Patients.

Supplement S3. Laboratory Parameters for BC(-) Sepsis Patients.

Supplement S4. Laboratory Parameters for BC(+) SIRS Patients.

Supplement S5. Laboratory Parameters for BC(-) SIRS Patients.

Supplement S6. BC(+) Sepsis Patients with the Highest Observed SeptiScores.

Supplement S7. BC(+) Sepsis Patients with Band 2 SeptiScores.

Supplement S8. BC(-) Sepsis Patients with Extreme Band 4 SeptiScores.

Supplement S9. BC(-) Sepsis Patients with Band 1 SeptiScores.

Supplement S10. BC(+) SIRS Patients.

Supplement S11. BC(-) SIRS Patients with Band 4 SeptiScores.

Supplement S12. Distribution of Patients According to Clinical Study, SeptiScore Band, Blood Culture Status, Sepsis / SIRS Diagnosis, and Likelihood Ratio (LR).

Supplement S13. Combinations of Idylla instrument software version, SeptiCyte RAPID test software version, and SeptiCyte RAPID cartridge lots that were used in this study.

**Abbreviations used in the Supplements:**

| Abbreviation | Meaning |
|---|---|
| AUC | area under ROC curve |
| BC | blood culture |
| CRP | C-reactive protein |
| CSF | cerebro-spinal fluid |
| HIV | human immunodeficiency virus |
| ICU | intensive care unit |
| IQR | inter-quartile range |
| KNST | Klebsiella-like non-spore-forming bacteria |
| LR | likelihood ratio |
| MAP | mean arterial pressure |
| Max | maximum |
| Min | minimum |
| NA | not available |
| ND | not determined |
| PCR | polymerase chain reaction |
| qSOFA | quick SOFA (score) |
| ROC | receiver operating characteristic (curve) |
| RPD | retrospective physician diagnosis |
| SIRS | Systemic Inflammatory Response Syndrome |
| SOFA | Sequential Organ Failure Assessment (score) |
| WBC | white blood cell |

**Supplement S1: Aggregate Characteristics of Ghanaian SIRS Patients.** The Ugandan SIRS patient set was supplemented with five SIRS patients from Ghana. Only the aggregate patient characteristics, not the individualized characteristics, were available for the 5 Ghanaian patients.

| Patient Characteristic | Median value (range) |
|---|---|
| Age (years) | 63 (39-78) |
| Sex distribution | 2 female, 3 male |
| 0 hr heart rate (beats/min) | 99 (71-117) |
| 0 hr MAP (mm Hg) | 107 (83-120) |
| 0 hr modified SOFA | 2 (0-5) |
| 0 hr lactate (mmol/L) | 2.47 (1.85-2.93) |
| 0 hr WBC (cells/uL) | 8,300 (5,700-18,500) |
| Days hospitalized | 11 (2-15) |

**Supplement S2. Laboratory Parameters for BC(+) Sepsis Patients.**

| | **Clinical Study** | | |
|---|---|---|---|
| **Patient characteristic** | **510k** | **UGANDA** | **SeptAsTERS** |
| BC(+) and called Sepsis | 48 | 22 | 15 |
| Gram (+) | 19 bacterial, 1 fungal | 10 bacterial, 1 fungal | 4 bacterial, 2 fungal |
| Gram (-) | 22 bacterial | 11 bacterial | 9 bacterial |
| Mixed Gram (+/-) | 5 bacterial, 1 bacterial + fungal | 0 | 0 |
| SeptiScore, median (range) | 8.2 (5.5-14.8) | 10.3 (6-15) | 9.4 (5.6-11.5) |
| Lactate (mmol/L), median (range) | 2.8 (0.5-15.7) [missing: 10] | NA | 2.95 (0.9-13.9) |
| CRP (mg/L), median (range) | 29 (6-291) [missing: 37] | 197.5 (38.4-200) [missing: 16] | 225 (38-353) |
| WBC (cells/uL), median (range) | Min: 12,500 (300-51,800)<br>Max: 17,600 (3,700-56,900)<br>[missing 1 value each] | 7,120 (2,130-17,200) [missing: 2] | 10,910 (1300-62,760) |

**Supplement S3. Laboratory Parameters for BC(-) Sepsis Patients.**

| | Clinical Study | | |
|---|---|---|---|
| **Patient characteristic** | **510k** | **UGANDA** | **SeptAsTERS** |
| BC(-) and called Sepsis | 33 | 112 | 17 |
| SeptiScore, median (range) | 7.8 (3.9-12.1) | 7.6 (2.7-13.1) | 6.9 (4-11.1) |
| Lactate (mmol/L), median (range) | 2.2 (0.7-13.3) | NA | 1.12 (0.68-7.46) |
| CRP (mg/L), median (range) | 15 (1-240) [missing: 26] | 99 (10-200) [missing: 101] | 127 (7-411) |
| WBC (cells/uL), median (range) | Max: 15,100 (1000-40,600) [missing: 2]<br>Min: 11,400 (300-37,200) [missing: 1] | 5,500 (1,970-74,600) [missing: 7] | 16,000 (7,000-32,000) |

**Supplement S4. Laboratory Parameters for BC(+) SIRS Patients.**

| | **Clinical Study** | | |
|---|---|---|---|
| **Patient characteristic** | **510k** | **UGANDA** | **SeptAsTERS** |
| BC(+) and called SIRS | 3 | 0 | 2 |
| SeptiScore, median (range) | 5.4 (4.9-5.4) | NA | 8.4 (7.4-9.4) |
| Lactate (mmol/L), median (range) | 1 [missing: 2 patients] | NA | 0.82 (0.69-0.94) |
| CRP (mg/L), median (range) | ND | NA | 213 (117-310) |
| WBC (cells/uL), median (range) | Min: 4,900 (4,600-11,400)<br>Max: 7,200 (4,900-12,100) | NA | 8,000 (4,000-11,000) |

**Supplement S5. Laboratory Parameters for BC(-) SIRS Patients.**

| | Clinical Study | | |
|---|---|---|---|
| **Patient characteristic** | **510k** | **UGANDA** | **SeptAsTERS** |
| BC(-) and called SIRS | 221 | 3 | 23 |
| SeptiScore, median (range) | 5.3 (1.8-11.3) | 8.9 (7.8-10.7) | 6 (3.2-11.3) |
| Lactate (mmol/L), median (range) | 2.1 (0.5-19) [missing: 84] | NA | 1.1 (0.54-13.19) |
| CRP (mg/L), median (range) | 5.0 (1-27) [missing: 212] | 156, 200 [missing: 1] | 127 (14-400) |
| WBC (cells/uL), median (range) | Min: 10,300 (400-29,000) [missing: 6]<br>Max: 12,000 (400-35,040) [missing: 8] | 7000, 12,750 [missing: 1] | 14,000 (7,000-29,000) |

**Supplement S6: BC(+) sepsis patients with the highest observed SeptiScores.**

This group of five BC(+) sepsis patients with the highest observed SeptiScores (12.9-15.0) consisted of male patients with ages ranging from 30 to 80 years. Four out of five patients were from the Uganda cohort, and one was from the 510k cohort. All five patients presented with severe infections that progressed to sepsis or septic shock, as indicated by clinical diagnoses. Severity scores were elevated (SOFA 5-7, qSOFA 1-2).

The types of primary infections varied: three patients were diagnosed with pneumonia, one with a complex *Staphylococcus aureus* infection, and one with acute gastroenteritis. Bacterial pathogens were confirmed in the bloodstream for all patients: *Streptococcus pneumoniae* was found in three patients, *Klebsiella pneumoniae* in one, and Methicillin-Sensitive *Staphylococcus aureus* (MSSA) in one.

Two of the patients from the Uganda cohort were HIV positive and on antiretroviral therapy.
All patients received intravenous antibiotics.

The clinical outcomes were diverse: two patients were discharged (after 5 and 10 days respectively), one patient died (after an 8-day hospital stay), one patient had a very brief hospital stay of 1 day (specific outcome like discharge or death not detailed), and one patient (age range 75-80) experienced a prolonged Intensive Care Unit (ICU) stay of 28 days.

**Supplement S7: BC(+) Sepsis Patients with Band 2 SeptiScores.**

This group of eight BC(+) sepsis patients consisted of five from the 510k cohort, two from the SeptAsTERS cohort, and one from the Uganda cohort. Sepsis was diagnosed by the care team for the Ugandan patient, and by RPD for the patients from the 510k and SeptsTERS cohorts. SeptiScores ranged from 5.5 to 6.0.

The patient from Uganda was HIV(+) and on antiretroviral therapy.

Primary sites of infection were identified initially or during hospital stay as abdominal, cellulitis, pulmonary, urinary tract, or kidney.

Pathogens with short times to BC positivity (within 1 day) were *Streptococcus anginosus,* and *Escherichia coli.* Pathogens with moderately long times to BC positivity (3-4 days) were *Staphylococcus aureus, Clostridioides difficile, Klebsiella pneumoniae; Corynebacterium spp., Staphylococcus epidermidis.* Pathogens with very long times to BC positivity (9-10 days) were *Fusobacterium, Propionobacterium spp., Parvimonas micra*.

All patients received intravenous antibiotics.

**Supplement S8. BC(-) Sepsis Patients with Extreme Band 4 SeptiScores.**

A group of five BC(-) sepsis patients with the highest observed SeptiScores (range 12.1-13.1) was considered. This group consisted of two male and three female patients, with ages ranging from 35 to 85 years. Four out of five patients were from the Uganda cohort, and one was from the 510k cohort. All five patients were diagnosed as septic either by the care team (Uganda) or by RPD (510k). Severity scores, when measured, were elevated (SOFA 1-5, qSOFA 1-3). None of the patients had septic shock.

The types of primary infections varied: three patients were diagnosed with pneumonia (COVID positive in two cases), one was diagnosed with UTI + gastroenteritis, and one with meningococcal meningitis. The patient with meningococcus meningitis also tested positive for EBV.

One patient was HIV (+) and on antiretroviral therapy.

Blood cultures, when taken, remained negative after 5 days incubation. However, all patients received intravenous antibiotics.

Hospital length of stay was 2-6 days. All patients survived.

**Supplement S9: BC(-) Sepsis Patients with Band 1 SeptiScores.**

Ten patients fell in this category, represented by the blue points in the Band 1 region of Figure 3B. There were four from the Uganda cohort, three from the 510k cohort, and three from the SeptAsTERS cohort. There were 5 females and 5 males, with age spanning the 15-70 year range.

SOFA scores ranged from 2 to 12, qSOFA scores ranged from 0 to 2. SeptiScores ranged from 2.7 to 4.7.

Primary sites of infection were identified for all ten patients, and were determined to be pulmonary, abdominal, peritonitis, hematological (malarial), pancreatic, urinary in origin.

Bacteria isolated from non-blood sites included: *Enterobacter cloacae*, *Eschericia coli*, *Entercoccus casseliflavus*, *Eschericia faecalis, Prevotella buccae*, *Raoutella ornithinolytica* (drainage fluid); *Enterococcus faecalis* (urine); *S. aureus* & *Haemophilus influenzae* (sputum). One patient tested positive in blood culture, but late after inclusion in the study (*Staphylococcus saccharolyticus)*. In addition, three patients from the Uganda cohort tested positive for malaria parasites in the blood (*Plasmodium falciparum)*.

All patients received intravenous antibiotics.

Three patients had prolonged hospital lengths of stay (> 28 days) and one patient died.

**Supplement S10: BC(+) SIRS Patients.**

This group of BC(+) SIRS patients are represented by the red points in Figure 3C. They consisted of three female and two male patients, with ages ranging from 45 to 80 years. Three of the patients were from the 510k cohort, and two were from the Heidelberg cohort. All patients were diagnosed with SIRS by RPD. SeptiScores ranged from 4.9 to 9.4.

Three of the patients were found to have true bloodstream infections. The site of primary infection was identified as abdominal in two cases, but was not identified in the third case. For two of the patients, *Bacillus spp. and Staphylococcus epidermidis* were identified from blood culture, and one patient also had a positive urine culture for *Enterococcus faecium*. The third patient had a positive blood culture for *Candida albicans,* as well as drain fluid positivity for *Klebsiella aerogenes, Candida albicans, Escherichia coli, Vancomycin-resistant Enterococcus, and Morganella morganii.* These three patients were treated with intravenous antibiotics, and the patient with the candidemia was also treated intravenously with an antifungal agent.

BC positive results were also obtained for the remaining two patients, but only after very long incubation (6-8 days), and no primary site of infection was identified. As these BC results were considered contaminants, the patients were not treated with antibiotics.

**Supplement S11: BC(-) SIRS Patients with Band 4 SeptiScores.**

A random selection of fourteen patients from the Band 4 region of Figure 3C was considered.
The following characteristics were noted:

9/14 (64%) male.

7/14 (50%) from 510k, 3/14 (21%) from Uganda, 4/14 (29%) from SeptAsters. (The study as a whole has 61% 510k patients, 27% Uganda patients, 12% SeptAsTERS patients. A chi square proportion test gives c2 = 3.58, p = 0.17 so there is no significant selection bias in the composition of patients in Supplement S11, relative to the proportions represented in the study as a whole.)

Age range 30-85 years
SOFA range 1-5, qSOFA range 0-2
SeptiScore range 7.4-11.3

primary site of infection: unidentified 10/14 (71%), abdominal 3/14 (21%), urinary 1/14 (7%)

Microbiology test results:

4 patients: cultures ordered, results negative

4 patients: either cultures not ordered, or results negative (510k cohort)

2 patients (combined data): Intraoperative smear: *Proteus mirabilis, Proteus hauseri, Enterococcus faecium, Streptococcus anginosus, Haemophilus parainfluenzae, Prevotella melaninogenica, Prevotella buccae, KNST [Klebsiella-like non-spore-forming bacteria]*

1 patient: drainage fluid: *Streptococcus anginosus, Streptococcus constellatus, Neisseria spp.*

1 patient: urinary catheter: *Escherichia coli*; vaginal swab: *Escherichia coli, Enterococcus spp.*

1 patient: Epstein-Barr virus (blood + CSF by PCR)

1 patient: skin swab: *Klebsiella pneumoniae, Proteus mirabilis*

All patients received intravenous antibiotics.

Hospital length of stay 1-19 days (IQR 4-9 days).

No patients died.

**Supplement S12. Distribution of Patients According to Clinical Study, SeptiScore Band, Blood Culture Status, Sepsis / SIRS Diagnosis, and Likelihood Ratio (LR).** Calculations based on 85 patients that were blood culture positive and-diagnosed with sepsis, and 257 patients that were diagnosed with SIRS (including 3 SIRS patients with positive blood cultures). Patients with a SeptiScore falling between 7.4 and 15 (Band 4) had a LR of 5.88 for BC(+) sepsis. Patients with a SeptiScore falling between 0 and 4.9 (Band 1) had a LR of 0 for BC(+) sepsis. Details of LR calculations are as follows. Point bi-serial correlations were calculated using the R package 'ltm' (Rizopoulos, D., *J. Stat. Softw*. **2006**, 17, 1–25).

- Band 1: BC(+) sepsis = 0, SIRS = 95, total = 95, Alpha = BC(+) sepsis in Band 1/total BC(+) sepsis= 0/85 = 0; Beta = SIRS in Band 1/total number of SIRS subjects = 95/257 = 0.37; LR (BC(+) sepsis / SIRS) = alpha/beta = 0/0.37 = 0.
- Band 2: BC(+) sepsis = 8, SIRS = 79, total = 87, Alpha = BC(+) sepsis in Band 2/total BC(+) sepsis= 8/85 = 0.094; Beta = 78/257 = 0.303; LR (BC(+) sepsis / SIRS) = 0.094/0.303 = 0.31. Point-biserial correlation SeptiScore vs. BC(+) = 0.2.
- Band 3: BC(+) sepsis = 11, SIRS = 51, total = 62, Alpha = BC(+) sepsis in Band 3/total BC(+) sepsis= 11/85 = 0.129; Beta = 50/257 = 0.195; LR (BC(+) sepsis / SIRS) = 0.129/0.195 = 0.66. Point-biserial correlation SeptiScore vs. BC(+) = 0.21.
- Band 4: BC(+) sepsis = 66, SIRS = 37, total = 103, Alpha = BC(+) sepsis in Band 4/total BC(+) sepsis= 66/85 = 0.776; Beta = 34/257 = 0.132; LR (BC(+) sepsis / SIRS) = 0.776/0.132 = 5.88. Point-biserial correlation SeptiScore vs. BC(+) =0.39.
- Point-biserial correlation SeptiScore vs. BC(+) (all data) = 0.65 (ROC AUC=0.91).

| SeptiCyte RAPID Band | 510k BC(+) Sepsis | 510k SIRS | Uganda BC(+) Sepsis | Uganda SIRS | SeptAsTERS BC(+) Sepsis | SeptAsTERS SIRS | Total BC(+) Sepsis | Total SIRS | Total Patients | LR |
|---|---|---|---|---|---|---|---|---|---|---|
| Band 4 (7.4 – 15) | 34 | 23 | 20 | 5 | 12 | 6 | 66 | 34 | 100 | 5.88 |
| Band 3 (6.2 – 7.3) | 9 | 41 | 1 | 2 | 1 | 7 | 11 | 50 | 61 | 0.66 |
| Band 2 (5.0 – 6.1) | 5 | 73 | 1 | 1 | 2 | 4 | 8 | 78 | 86 | 0.31 |
| Band 1 (0 – 4.9) | 0 | 87 | 0 | 0 | 0 | 8 | 0 | 95 | 95 | 0 |
| **Total** | **48** | **224** | **22** | **8** | **15** | **25** | **85** | **257** | **348** | |

**Supplement S13. Combinations of Idylla instrument software version, SeptiCyte RAPID test type and software version, and SeptiCyte RAPID cartridge lots used in this study.**

| Sample Source | Instrument Software version | SeptiCyte RAPID test type | SeptiCyte RAPID software version | SeptiCyte RAPID cartridge lot number |
|---|---|---|---|---|
| Uganda | 26.0 | RUO 1.0 | 1.14 | 5341 |
| Uganda | 26.0 | IVD 1.20 | 1.23 | 5621 |
| Uganda | 27.0 | IVD 1.20 | 1.23 | 5815 |
| Uganda | 27.0 | IVD 1.20 | 1.23 | 6102 |
| Uganda | 27.0 | IVD 2.20 | 2.25 | 6448 |
| Uganda | 27.0 | IVD 2.20 | 2.25 | 6530 |
| Ghana | 27.0 | IVD 2.20 | 2.25 | 6530 |
| 510k | 26.0 | IUO 1.0 | 1.30 | 4613 |
| 510k | 26.0 | IUO 1.0 | 1.30 | 4614 |
| Heidelberg | 26.0 | IVD 2.1 | 2.5 | 5977 |
